# Communicating risk to patients: developing an “Efficiency Index” for diagnostic and screening tests

**DOI:** 10.1101/2021.08.18.21262205

**Authors:** Andrew J Larner

## Abstract

Diagnostic and screening tests may have risks such as misdiagnosis, as well as the potential benefits of correct diagnosis. Effective communication of this risk to both clinicians and patients can be problematic. The purpose of this study was to develop a metric called the “efficiency index” (EI), defined as the ratio of test accuracy and inaccuracy, or the number needed to misdiagnose divided by number needed to diagnose. This was compared with a previously described “likelihood to be diagnosed or misdiagnosed” (LDM), also based on “numbers needed” metrics. Datasets from prospective pragmatic test accuracy studies examining four brief cognitive screening instruments (Mini-Mental State Examination; Montreal Cognitive Assessment; Mini-Addenbrooke’s Cognitive Examination, MACE; and Free-Cog) were analysed to calculate values for EI and LDM, and to examine their variation with test cut-off for MACE. EI values were also calculated using a modification of McGee’s heuristic for the simplification of likelihood ratios to estimate percentage change in diagnostic probability. The findings indicate that EI is easier to calculate than LDM and, unlike LDM, may be classified either qualitatively or quantitatively in a manner similar to likelihood ratios. EI shows the utility or inutility of diagnostic and screening tests, illustrating the inevitable trade-off between diagnosis and misdiagnosis. It may be a useful metric to communicate risk in a way that is easily intelligible for both clinicians and patients.

## Introduction

No medical treatment or test is without potential harms as well as benefits, and hence associated with risk. Communicating such risk to patients for the purpose of shared decision making has attracted much attention and research in recent times, for example into the most appropriate methods by which to achieve such communication effectively. Guidance on both verbal and numerical qualifiers of risk has appeared.^1,2^ As regards numerical qualifiers, options often used in the context of therapeutic interventions include absolute risk (AR), relative risk (RR), and the number needed to treat (NNT),^3^ but no consensus on the optimum method has been established.

Considering diagnostic or screening tests, performance is typically described by comparison with a reference standard, such as a criterion diagnosis or a reference test, by constructing a 2×2 contingency table, such that all (N) index test results may be cross-tabulated as true positive (TP), false positive (FP), false negative (FN), or true negative (TN). From this standard 2×2 contingency table (Figure 1), various parameters of test discrimination may be calculated, many of which are familiar to clinicians as descriptors of test performance, such as sensitivity (Sens; or true positive rate) and specificity (Spec; or true negative rate), positive and negative predictive values, and positive and negative likelihood ratios (LR+, LR-).^4^

**Figure 1:**
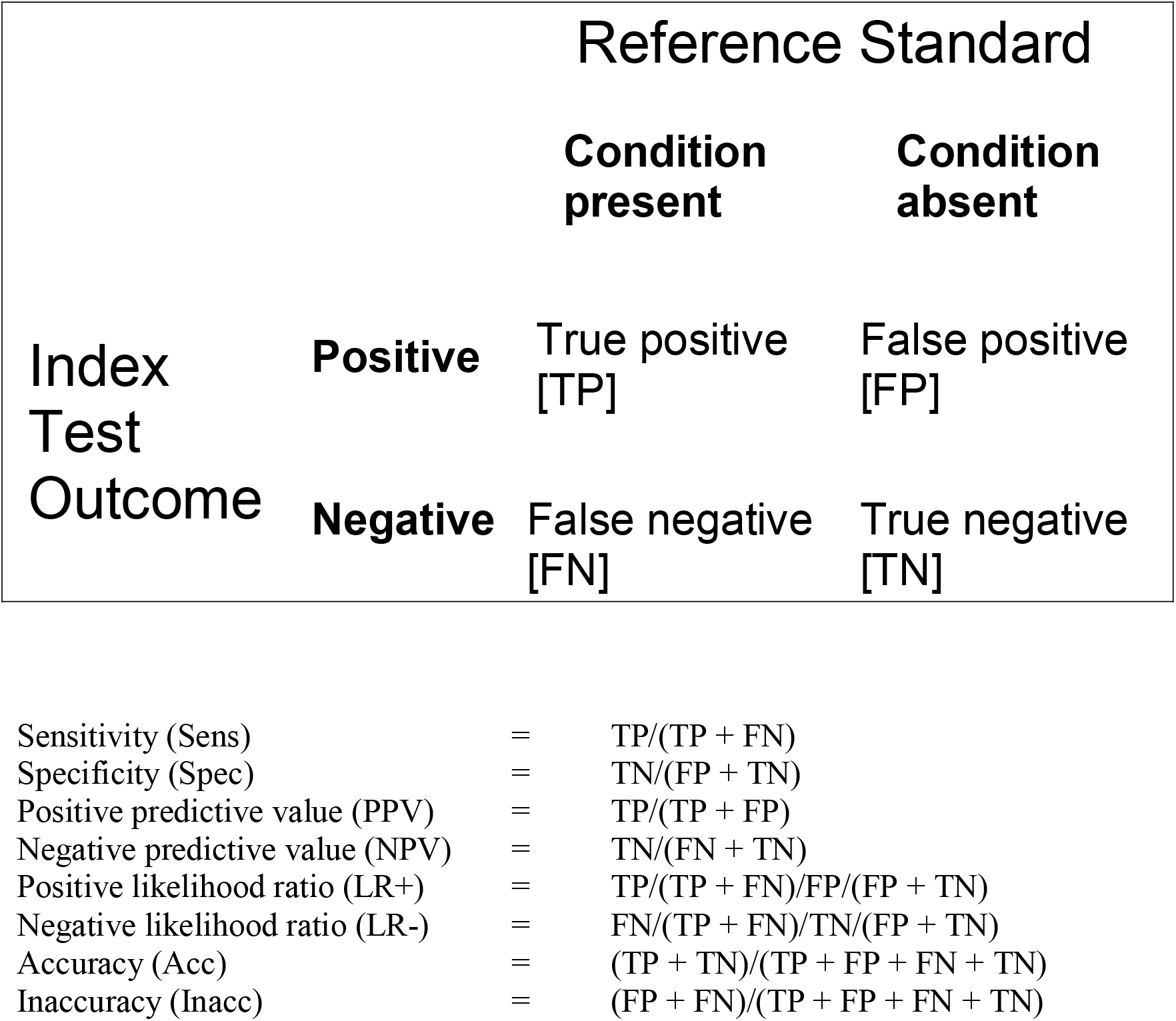
Standard 2×2 contingency table for diagnostic or screening test accuracy studies and formulae for paired measures.

One may thus distinguish from the 2×2 contingency table two conditions or relations between the index test and the reference standard: consistency, or matching, of outcomes (+/+ or TP, and -/- or TN); and contradiction, or mismatching (+/- or FP, and -/+ or FN). From these two conditions, the paired complementary parameters of accuracy (Acc = TP+TN/N) and inaccuracy or error rate (Inacc = FP+FN/N) may be derived. As negations, these may also be described using the Boolean NOT operator, since Acc = 1 – Inacc and Inacc = 1 – Acc.

How might these various test measures be effectively communicated to patients who are unfamiliar with the principles and nomenclature of binary classicism? A metric called the “likelihood to be diagnosed or misdiagnosed” (LDM) has been developed^5,6^ which may be useful for the purpose of communicating risk, specifically the risk of testing leading to misdiagnosis as opposed to correct diagnosis. LDM was conceptualised as analogous to the “likelihood to be helped or harmed” (LHH) metric which was developed to communicate the results of therapeutic (randomised controlled) trials. LHH is based on “number needed to” metrics, specifically the number needed to treat (NNT) for a specified treatment outcome (e.g. cure, remission, 50% reduction in symptoms)^7^ and the number needed to harm (NNH).^8^ LHH is the ratio of NNH to NNT, which is desirably as large as possible (high NNH, low NNT), thus summarising treatment benefits and risks.^9^

For diagnostic and screening tests one may, analogously, combine the number needed to diagnose (NND), as defined by Linn and Grunau as 1/(Sens + Spec – 1) or 1/Youden index (Y),^10^ and the number needed to misdiagnose (NNM), as defined by Habibzadeh and Yadollahie, as 1/Inacc,^11^ to produce a “likelihood to be diagnosed or misdiagnosed” (LDM) metric, where LDM = NNM/NND, which is desirably as large as possible (high NNM, low NND),^5,6^ thus summarising testing benefits and risks.

The LDM metric has proved serviceable in evaluating a wide range of neurological signs and cognitive screening instruments (CSIs) used in the evaluation of disorders of cognition.^5,6,12^ Nevertheless, LDM has some limitations and shortcomings. Consistent with its ad hoc development, based on existing metrics, LDM combined rates with different denominators which are not easily reconciled. Calculation of several parameters from the 2×2 table is required to reach LDM (Sens, Spec, Y, NND, Inacc, NNM), although online calculators exist.^13^ Furthermore, the “number needed to diagnose” based on the Youden index incorporates considerations not only of diagnosis but also of misdiagnosis, since Sens = 1 – false negative rate, and Spec = 1 – false positive rate. The resulting LDM has boundary values of -1 (useless test: Sens = Spec = 0, NND = -1; Inacc = 1, NNM = 1) and ∞ (perfect test: Sens = Spec = 1, NND = 1; Inacc = 0, NNM = ∞), and so the LDM values cannot be perfectly equated with the qualitative classification scheme developed for likelihood ratios (LR),^14^ which has boundary values of 0 and ∞, although LDM shares with LR an inflection point at 1 (LDM < 1 favours misdiagnosis, LDM > 1 favours diagnosis).^5,6^

A simple method to overcome these shortcomings of LDM may be proposed. The “number needed to diagnose” may be redefined, as NND*, using Acc, rather than Sens and Spec, such that NND* = 1/Acc. This formulation is analogous to the previous definition of number needed to misdiagnose,^11^ where NNM = 1/Inacc. Both these measures now share the same denominator from the 2×2 contingency table, N, and calculation is thus simplified, such that:

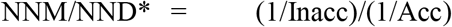

Whilst this ratio might justifiably be termed a “likelihood to be diagnosed or misdiagnosed”, an alternative name would be preferable to avoid confusion with the previously defined LDM. Kraemer denoted TP+TN as “efficiency”,^15^ so FP+FN might be termed “inefficiency”, and hence the ratio of efficiency/inefficiency may be denoted as the “efficiency index” (EI). Hence:

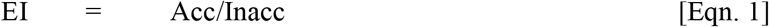

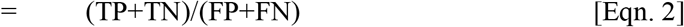

The boundary values of EI are 0 (useless test: Acc = 0; Inacc = 1) and ∞ (perfect test: Acc = 1, Inacc = 0), as for likelihood ratios.

The primary aim of this study was to examine the utility of the efficiency index and compare it to the previously defined LDM parameter when applied to test accuracy studies of several brief CSIs, namely the Mini-Mental State Examination (MMSE),^16^ the Montreal Cognitive Assessment (MoCA),^17^ the Mini-Addenbrooke’s Cognitive Examination (MACE),^18^ and the Free-Cog.^19^ Secondary aims were to examine other methods to calculate EI and to compare performance with LRs, particularly with McGee’s method of simplifying LR values as percentage changes in diagnostic probability.^20^

## Methods

Data from pragmatic prospective screening test accuracy studies using a standardised methodology were re-analysed. The studies were undertaken in a dedicated cognitive disorders clinic located in a secondary care setting (regional neuroscience centre) and examined four brief cognitive screening instruments (administration time ca. 5-10 min), all with a denominator of 30 points: Mini-Mental State Examination (MMSE),^21,22^ Montreal Cognitive Assessment (MoCA),^23^ Mini-Addenbrooke’s Cognitive Examination (MACE),^24^ and Free-Cog.^25^

In each study, criterion diagnosis of dementia followed standard diagnostic criteria (DSM-IV) and was made independent of scores on index CSIs to avoid review bias. For each study, prevalence of dementia was calculated as the sum of TP and FN divided by the total number of patients (N) assessed. All studies followed either the STAndards for the Reporting of Diagnostic accuracy studies (STARD)^26^ or the derived guidelines specific for dementia studies, STARDdem,^27^ dependent on the exact date at which each study was undertaken. In all studies subjects gave informed consent and study protocols were approved by the institute’s committee on human research (Walton Centre for Neurology and Neurosurgery Approval: N 310).

For each CSI the following parameters were calculated: Acc, Inacc, Youden index, LDM; and efficiency index by using the values of Acc and Inacc (Eqn. 1). A previously described measure based on Acc and Inacc, the identification index (II),^28^ was also calculated, defined as II = (Acc – Inacc) = 2.Acc – 1.

The variation of EI with test cut-off was examined using data from the test accuracy study of MACE^24^ and compared with the variation in LDM. EI was also calculated from values of test Sens and Spec in the MACE study. Since:

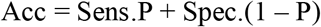

where P = prevalence of dementia; and

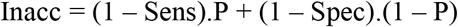

Hence:

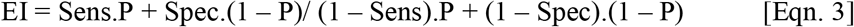

The performance of EI was also compared to that of LRs which may be used to calculate difference in pre- and post-test odds, since post-test odds = pre-test odds x LR. McGee showed that LR+ values of 2, 5, and 10 increased the probability of diagnosis by approximately 15%, 30%, and 45% respectively, whereas LR-values of 0.5, 0.2, and 0.1 decreased the probability of diagnosis by approximately 15%, 30% and 45% respectively. These figures derive from the almost linear relationship of probability and the natural logarithm of odds over the range 0.1 to 0.9, such that the percentage change in probability may be calculated independent of pre-test probability as:

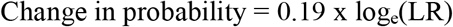

This simple heuristic obviates calculations between pre- and post-test odds and probabilities.^20^ As the boundary values of EI (0, ∞) correspond to those of LRs, calculations were undertaken to assess whether or not the heuristic described for LR values also holds for EI values. These calculations used data from the studies of MACE^24^ and MoCA.^23^ As both of these studies had a similar (low) pre-test probability of dementia, the issue was further examined using data from a test accuracy study of the Test Your Memory (TYM) test,^29^ this being the study with the highest pre-test probability of dementia reported from this clinic.^30^

## Results

A summary of the studies of MMSE, MoCA, MACE, and Free-Cog (Table 1) showed broadly similar prevalence of dementia, median age, and gender ratio in each patient cohort.

**Table 1:** Study demographics and test thresholds for dementia.

| **CSI** | **N** | **P**<br>**= prevalence**<br>**of dementia**<br>**= (TP + FN)/N** | **Age,**<br>**median**<br>**(years)** | **Gender**<br>**(F:M; %F)** | **Test threshold**<br>**for dementia** | **Ref(s)** |
| --- | --- | --- | --- | --- | --- | --- |
| MMSE | 244 | 0.18 | 60 | 117:127; 48 | <26/30 | 21,22 |
| MoCA | 260 | 0.17 | 59 | 118:142; 45 | <26/30 | 23 |
| MACE | 755 | 0.15 | 60 | 352:403; 47 | ≤20/30 | 24 |
| Free-Cog | 141 | 0.11 | 62 | 61:80, 43 | ≤22/30 | 25 |

Abbreviations: CSI = Cognitive screening instrument; TP = true positive; FN = false negative; MMSE = Mini-Mental State Examination; MoCA = Montreal Cognitive Assessment; MACE = Mini-Addenbrooke's Cognitive Examination

Comparing the various metrics for the diagnosis of dementia vs no dementia for each of these CSIs (Table 2) showed a similar ranking (best to worst) for Acc, Y, LDM, EI, and II.

**Table 2:** Comparing metrics for diagnosis of dementia vs no dementia by CSI (using cut-offs in Table 1)

| **CSI** | **Acc** | **Inacc** | **Y**<br>(= Sens + Spec – 1) | **LDM**<br>(= NNM/NND) | **EI**<br>(= Acc/Inacc) | **II**<br>(= 2.Acc – 1) |
| --- | --- | --- | --- | --- | --- | --- |
| MMSE | 0.676 | 0.324 | 0.497 | 1.536 | 2.089 | 0.352 |
| MoCA | 0.427 | 0.573 | 0.313 | 0.547 | 0.745 | -0.146 |
| MACE | 0.738 | 0.262 | 0.619 | 2.360 | 2.817 | 0.475 |
| Free-Cog | 0.709 | 0.291 | 0.670 | 2.320 | 2.439 | 0.418 |

Abbreviations: CSI = Cognitive screening instrument; Acc = correct classification accuracy; Inacc = Inaccuracy; II = Identification index; Y = Youden index; LDM = likelihood to be diagnosed or misdiagnosed; NNM = number needed to misdiagnosis; NND = number needed to diagnose; EI = Efficiency index; MMSE = Mini-Mental State Examination; MoCA = Montreal Cognitive Assessment; MACE = Mini-Addenbrooke's Cognitive Examination

Comparing the EI and LDM metrics across a range of MACE cut-offs (Table 3, Figure 2) showed that, as for the other CSIs (Table 2), EI was a more optimistic score than LDM and that, unlike II, EI nowhere had a negative value. The maxima for EI and LDM almost coincided (LDM ≤15/30; EI ≤14/30). Values for EI and LDM were approximately equal at higher test cut-offs but diverged at lower cut-offs, which may be a reflection of high sensitivity and low specificity of MACE for the diagnosis of dementia.^24^

**Table 3:** Diagnosis of dementia: comparing metrics at various MACE cut-offs.

| **Cut-off** | **Acc** | **Inacc** | **Y** | **LDM** | **EI** | **II** |
| --- | --- | --- | --- | --- | --- | --- |
| ≤29/30 | 0.170 | 0.830 | 0.02 | 0.02 | 0.204 | -0.66 |
| ≤28/30 | 0.197 | 0.803 | 0.05 | 0.06 | 0.246 | -0.61 |
| ≤27/30 | 0.262 | 0.738 | 0.12 | 0.16 | 0.355 | -0.48 |
| ≤26/30 | 0.336 | 0.664 | 0.21 | 0.33 | 0.507 | -0.33 |
| ≤25/30 | 0.417 | 0.583 | 0.31 | 0.53 | 0.716 | -0.17 |
| ≤24/30 | 0.495 | 0.505 | 0.39 | 0.76 | 0.982 | -0.01 |
| ≤23/30 | 0.560 | 0.440 | 0.47 | 1.07 | 1.27 | 0.12 |
| ≤22/30 | 0.625 | 0.375 | 0.53 | 1.43 | 1.67 | 0.25 |
| ≤21/30 | 0.687 | 0.313 | 0.59 | 1.90 | 2.20 | 0.37 |
| ≤20/30 | 0.738 | 0.262 | 0.62 | 2.36 | 2.82 | 0.48 |
| ≤19/30 | 0.771 | 0.229 | 0.61 | 2.67 | 3.36 | 0.54 |
| ≤18/30 | 0.801 | 0.199 | 0.60 | 3.00 | 4.03 | 0.60 |
| ≤17/30 | 0.808 | 0.192 | 0.56 | 2.95 | 4.21 | 0.62 |
| ≤16/30 | 0.841 | 0.159 | 0.58 | 3.63 | 5.29 | 0.68 |
| ≤15/30 | 0.860 | 0.140 | 0.56 | 4.00 | 6.12 | 0.72 |
| ≤14/30 | 0.868 | 0.132 | 0.51 | 3.92 | 6.55 | 0.74 |
| ≤13/30 | 0.866 | 0.134 | 0.41 | 3.15 | 6.48 | 0.73 |
| ≤12/30 | 0.866 | 0.134 | 0.37 | 2.79 | 6.48 | 0.73 |

Abbreviations: CSI = Cognitive screening instrument; Acc = correct classification accuracy; Inacc = Inaccuracy; II = Identification index; Y = Youden index; LDM = likelihood to be diagnosed or misdiagnosed; NNM = number needed to misdiagnosis; NND = number needed to diagnose; EI = Efficiency index

**Figure 2:**
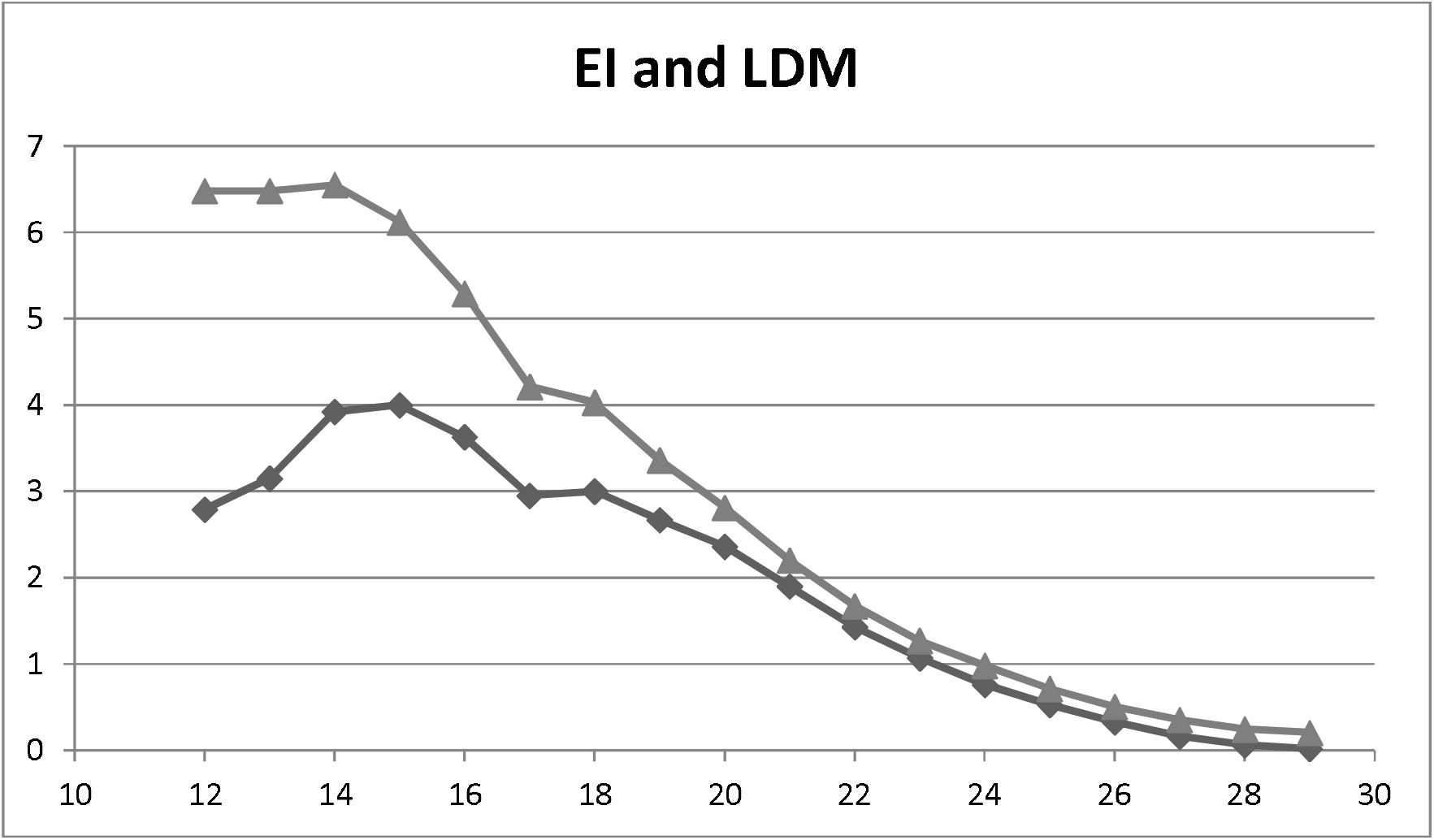
Plot of Efficiency Index (EI; upper line, triangles) and of Likelihood to be Diagnosed or Misdiagnosed (LDM; lower line, diamonds) values (y axis) vs MACE cut-off score (x-axis)

Using Eqn. 1, the value of EI for MACE was 2.817 (Table 2). This value was checked by using Eqn. 3, substituting the values for dementia prevalence (P = 0.151) and Sens and Spec at the test threshold for MACE (≤20/30; Table 1), respectively 0.912 and 0.707. Hence,

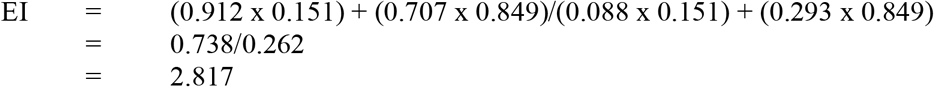

The same value of EI was thus obtained using two different methods.

To examine whether or not McGee’s simple rules obviating calculations between pre- and post-test odds and probabilities for LRs^20^ are also applicable to EIs, data from the MACE study were used,^24^ wherein:

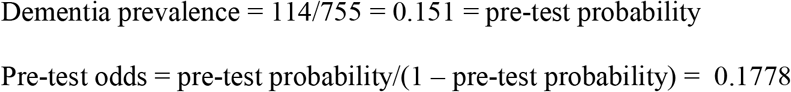

It is known that:

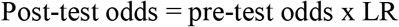

Substituting EI for LR, let:

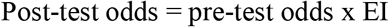

In the MACE study, EI = 2.817, favouring correct diagnosis. Hence:

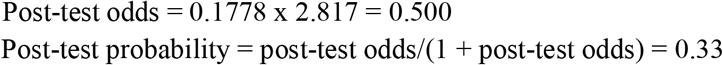

So, using calculations based on the observed pre-test probability, MACE increased diagnostic probability of dementia in this patient cohort from about 15% to about 33%, an approximately 18% increase.

Using the equation derived by McGee to calculate change in diagnostic probability independent of pre-test probability,^20^ and substituting LR with EI:

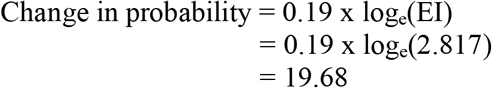

Thus similar values for percentage change in probability were obtained using two different methods.

Calculations of change in diagnostic probability were performed assuming EI values of 2.0, 5.0, 10.0, 0.5, 0.2, and 0.1 using both the methods, i.e. dependent and independent of observed pre-test probability. Similar calculations were also performed using the MoCA test accuracy study data,^23^ in which pre-test probability was similar to MACE but EI was 0.745 (i.e. favouring misdiagnosis). The results (Table 4) show that for EI values >1, favouring correct diagnosis, the percentage changes in diagnostic probability were similar when calculated independent of pre-test probability (column 1) and when calculated using observed pre-test probabilities (columns 2 and 3), approximating McGee’s 15, 30, and 45% increases for EI = 2, 5 and 10. However, for EI values <1, favouring misdiagnosis, McGee’s 15, 30, and 45% decreases for EI 0.5, 0.2 and 0.1 were not observed, presumably because of the low pre-test probabilities (ca. 15%) in these patient cohorts. In the S-shaped curve which describes the relationship between probability and log_e_ odds,^20^ this may correspond to the part of the plot away from the nearly linear portion which runs from about 0.1 to about 0.9.

**Table 4:** EI values calculated independent of pre-test probability and for selected cognitive screening instruments based on pre-test probability observed in test accuracy studies.

| **EI** | **% change in diagnostic probability calculated independent of pre-test probability as $0.19 \times \log_e(\text{EI})$** | **MACE: % change in diagnostic probability based on pre-test probability $(0.15)^{24}$** | **MoCA: % change in diagnostic probability based on pre-test probability $(0.17)^{23}$** | **TYM: % change in diagnostic probability based on pre-test probability $(0.35)^{30}$** |
| --- | --- | --- | --- | --- |
| 10.0 | +43.7 | +49 | +54 | +49 |
| 5.0 | +30.5 | +32 | +34 | +38 |
| 4.882 (TYM) | +30.1 | - | - | +37 |
| 2.817 (MACE) | +19.7 | +18 | - | - |
| 2.0 | +13.2 | +11 | +12 | +17 |
| 1.0 | 0 | 0 | 0 | 0 |
| 0.745 (MoCA) | -5.6 | - | -4 | - |
| 0.5 | -13.2 | -7 | -8 | -14 |
| 0.2 | -30.5 | -12 | -13 | -25 |
| 0.1 | -43.7 | -13 | -15 | -30 |

To further examine this point, data from a test accuracy study of the Test Your Memory (TYM) test^30^ were reanalysed, wherein the pre-test probability of dementia was 0.35, the highest reported in studies from this clinic. The change in probability based on the observed pre-test probability (Table 4, column 4) approximated the values calculated independent of pre-test probability (column 1) more closely than for MACE and MoCA for EI values <1.

## Discussion

This paper defines a new parameter, the efficiency index (EI), which may be of use not only for the evaluation of diagnostic and screening tests but also for the communication of risk to both clinicians and patients.

EI may be conceptualised, like a previously defined “likelihood to be diagnosed or misdiagnosed” (LDM) parameter, as a ratio of test harms (misdiagnosis) and benefits (diagnosis), and hence a measure of what has previously been termed the “fragility” of screening and diagnostic tests.^31^ Indeed, it might have been named the “fragility index”, understood as a propensity to break or fail. However, “efficiency index” emphasizes its relation to efficiency, understood as the ability to do things well, and conceptualised as a ratio of useful output to total input, or product per cost. However, EI differs from efficiency in that efficiency always has a value <1, whereas the upper bound of the EI, for a perfect diagnostic or screening test, is ∞.

For clinicians evaluating diagnostic or screening tests, EI has kinship with, but advantages over, LDM. It is more easily explicable (and, hence, more elegant) than the makeshift (although not arbitrary) derivation of LDM. EI is easier to calculate than LDM, requiring at its simplest only the four values from the cells of the 2×2 contingency table (Eqn. 2), whilst retaining the inflection point at the value of 1, as for LRs (EI > 1 indicates greater likelihood of correct diagnosis; EI < 1 indicates greater likelihood of misdiagnosis). Based on the calculations (Table 4), it seems appropriate to use the same classification scheme for EI values as proposed by Jaeschke et al. for LRs^14^ (Table 5, column 1), although it should be noted that this is a qualitative classification. McGee’s simplification of LRs also appears to be applicable to EI values (Table 4), and hence this numerical classification (percentage change in diagnostic probability) might also be used (Table 5, column 2). Whilst both LRs and EI can be calculated from the values of Sens and Spec, EI is dependent on prevalence (Eqn. 3) whereas LRs are not (at least algebraically, although in practice there may be variation).

**Table 5:** Suggested classification of EI values.

| **EI value** | **Qualitative classification of change in probability of diagnosis (after Jaeschke et al.<sup>14</sup>)** | **Approximate % change in probability of diagnosis (after McGee<sup>20</sup>)** |
| --- | --- | --- |
| $\leq 0.1$ | Very large decrease | - |
| 0.1 | Large decrease | -45 |
| 0.2 | Large decrease | -30 |
| 0.5 | Moderate decrease | -15 |
| 1.0 |  | 0 |
| 2.0 | Moderate increase | +15 |
| 5.0 | Moderate increase | +30 |
| 10.0 | Large increase | +45 |
| $\geq 10.0$ | Very large increase | - |

How does EI compare to other unitary measures which have been used to summarise diagnostic or screening test performance? Unlike the simple subtraction of Inacc from Acc, previously described as the identification index (II),^28^ EI does not produce negative values which have been previously noted to occur with II (if Inacc > Acc; Table 3) and whose meaning is difficult to comprehend (indeed may be meaningless).^24^ Part of the reason for this may be that, unlike II, EI is developed from “number needed to” metrics (NND*, NNM), whereas II was used as the basis for a “number needed to” metric, the “number needed to screen”.^28^

Other unitary measures which have been used to summarise diagnostic test performance, and with which EI should be compared, include the Youden index (Y), diagnostic odds ratio (DOR), critical success index, F measure, area under the receiver operating curve (AUC ROC), and Matthews’ correlation coefficient (MCC).^4,31,32^ Being based on Acc and Inacc, EI treats FN and FP as equally undesirable, as is also the case for Y and DOR, an assumption which is often not the case in clinical practice where FN may be considered more costly. As Acc is calculated using values from both columns of the 2×2 contingency table, EI is dependent on disease prevalence. The maximal value of Y arbitrarily assumes disease prevalence to be 50%. DOR gives the most optimistic results by choosing the best quality of a test and ignoring its weaknesses, particularly in populations with very high or very low risk. Ratios of DOR become unstable and inflated as denominator approaches zero, which is also true of EI. Critical success index, F measure, and AUC ROC all ignore TN values, unlike EI. AUC ROC combines test accuracy over a range of thresholds which may be both clinically relevant and clinically nonsensical. MCC takes into account the size of all four classes in the 2×2 contingency table and is widely regarded as being a very informative score for establishing the quality of a binary classifier, but the calculation (geometric mean of Y and the predictive summary index) is less straightforward than for EI.

For the communication of risk to patients, use of qualitative information is generally discouraged because of the potential ambiguity of such terms.^1,2^ Hence the suggested qualitative classification of EI values (Table 5, column 1) may not be of use in this situation. However, unlike other quantitative measures, EI involves no fractions, frequencies, or percentages, which may be advantageous when discussing risk with those with low numeracy skills.^1,2^ EI is based on “number needed” metrics which were originally deemed more intuitive for patients as well as clinicians,^7^ but empirical studies have suggested that NNT is more difficult for patients to understand than AR and RR.^3,33^

In summary, the proposed efficiency index may prove an acceptable unitary measure of diagnostic and screening test utility for clinicians as it is easy to calculate and interpret. It may also be useful for communicating risk of diagnosis and misdiagnosis to patients but further empirical studies will be required specifically to address this question.

## Data Availability

Available on any reasonable request

## Funding and Acknowledgements

None

